# Post-acute and Chronic Kidney Function Outcomes of COVID-19 in Children and Adolescents: An EHR Cohort Study from the RECOVER Initiative

**DOI:** 10.1101/2024.06.25.24309488

**Authors:** Lu Li, Ting Zhou, Yiwen Lu, Jiajie Chen, Yuqing Lei, Qiong Wu, Jonathan Arnold, Michael J. Becich, Yuriy Bisyuk, Saul Blecker, Elizabeth Chrischilles, Dimitri A Christakis, Carol Reynolds Geary, Ravi Jhaveri, Leslie Lenert, Mei Liu, Parsa Mirhaji, Hiroki Morizono, Abu Saleh Mohammad Mosa, Ali Mirza Onder, Ruby Patel, William E. Smoyer, Bradley W. Taylor, David A. Williams, Bradley P. Dixon, Joseph T Flynn, Caroline Gluck, Lyndsay A. Harshman, Mark M Mitsnefes, Zubin J. Modi, Cynthia G. Pan, Hiren P. Patel, Priya S. Verghese, Christopher B. Forrest, Michelle R. Denburg, Yong Chen, the RECOVER Consortium

## Abstract

We investigated the risks of post-acute and chronic adverse kidney outcomes of SARS-CoV-2 infection in the pediatric population via a retrospective cohort study using data from the RECOVER program. We included 1,864,637 children and adolescents under 21 from 19 children’s hospitals and health institutions in the US with at least six months of follow-up time between March 2020 and May 2023. We divided the patients into three strata: patients with pre-existing chronic kidney disease (CKD), patients with acute kidney injury (AKI) during the acute phase (within 28 days) of SARS-CoV-2 infection, and patients without pre-existing CKD or AKI. We defined a set of adverse kidney outcomes for each stratum and examined the outcomes within the post-acute and chronic phases after SARS-CoV-2 infection. In each stratum, compared with the non-infected group, patients with COVID-19 had a higher risk of adverse kidney outcomes. For patients without pre-existing CKD, there were increased risks of CKD stage 2+ (HR 1.20; 95% CI: 1.13-1.28) and CKD stage 3+ (HR 1.35; 95% CI: 1.15-1.59) during the post-acute phase (28 days to 365 days) after SARS-CoV-2 infection. Within the post-acute phase of SARS-CoV-2 infection, children and adolescents with pre-existing CKD and those who experienced AKI were at increased risk of progression to a composite outcome defined by at least 50% decline in estimated glomerular filtration rate (eGFR), eGFR <15 mL/min/1.73m^2^, End Stage Kidney Disease diagnosis, dialysis, or transplant.

**Lay abstract:** This study examined the impact of COVID-19 on kidney health in children and adolescents under 21 years old in the United States. Using data from the RECOVER program, we analyzed the health records of 1,864,637 young individuals from 19 hospitals and health institutions between March 2020 and May 2023. The study focused on three groups: those with pre-existing chronic kidney disease (CKD), those who experienced acute kidney injury (AKI) during the initial COVID-19 infection, and those without any prior kidney issues. The results showed that children and adolescents who had COVID-19 were at a higher risk of developing serious kidney problems later on, even if they had no previous kidney conditions. This research highlights the long-term effects of COVID-19 on kidney health in young people and underscores the importance of monitoring kidney function in pediatric COVID-19 patients.

## Introduction

Recent research has shed light on the post-acute sequelae of SARS-CoV-2 (PASC), commonly referred to as long COVID^1–4^, which can manifest beyond the initial four-week acute phase. While initially recognized predominantly among adults, the emergence of PASC has raised questions about its impact on the pediatric population. In the United States, PASC has affected 5-10% of children, which is comparable to adults (6.9%)^5,6^. However, there are important differences in the presentation and outcomes of acute SARS-CoV-2 infection between children and adults. Children can have different symptoms compared to adults and tend to have a milder disease course, with a lower risk of hospitalization and death, particularly among children without pre-existing conditions^7–9^. Given these differences in acute infection, as well as the differences in prevalence between children and adults, the characteristics of PASC require further study in children.

One notable study by Bowe et al., using national health databases from the US Department of Veterans Affairs, reported that after the first 30 days post-infection, individuals with COVID-19 exhibited increased risks of several adverse kidney outcomes: acute kidney injury (AKI); declines in estimated glomerular filtration rate (eGFR) of >=30%, >=40%, and >=50% from baseline; kidney replacement therapy (KRT); and major adverse kidney events (MAKE), defined as a composite of >=50% eGFR decline, KRT, or all-cause mortality^10^. Conversely, studies focusing on children and adolescents have been limited by short duration of follow-up, sample size (mostly less than 100 patients), and a narrow selection of outcomes: AKI, mortality, and multisystem inflammatory syndrome in children (MIS-C)^11–15^.

This paper aims to bridge these gaps in knowledge by examining a set of post-acute kidney outcomes among children and adolescents after SARS-CoV-2 infection. To assess the impact of PASC on pediatric patients with pre-existing kidney injury, we stratified all analyses on the presence of pre-existing CKD and AKI experienced during the acute phase of the infection. Our objective was to understand the risks of post-acute kidney manifestations of SARS-CoV-2 infection in the pediatric population, thereby informing future care strategies for those affected by long COVID. (hypothesis?)

## Methods

### Data Sources

This study is part of the NIH Researching COVID to Enhance Recovery (RECOVER) Initiative (https://recovercovid.org/), which aims to learn about the long-term effects of COVID-19. Nineteen sites contributed to the data: Cincinnati Children’s Hospital Medical Center, Children’s Hospital of Philadelphia, Children’s Hospital of Colorado, Duke University, University of Iowa Healthcare, Ann & Robert H. Lurie Children’s Hospital of Chicago, Medical College of Wisconsin, University of Michigan, University of Missouri, Children’s Hospital at Montefiore, Medical University of South Carolina, Children’s National Medical Center, Nationwide Children’s Hospital, University of Nebraska Medical Center, Nemours Children’s Health System (in Delaware and Florida), Northwestern University, New York University School of Medicine, OCHIN, Ohio State University, University of Pittsburgh, Seattle Children’s Hospital, Stanford Children’s Health, University of California San Francisco, Vanderbilt University Medical Center, Wake Forest University Health Sciences, Weill Cornell Medical College, Emory University, and Louisiana State University. Data was extracted from the RECOVER Database Version s9.

### Cohort Construction

We conducted a retrospective study spanning from March 2020 to May 2023, with a cohort entry date cutoff of December 2022 (179 days before the end of the study period). The index date for these patients was defined as the first indication of SARS-CoV-2 infection. We included patients under the age of 21, who had at least one visit within 24 months to 7 days prior to the index date (defined as the baseline period) and at least one encounter within 28 to 179 days after the index date (defined as the follow-up period). For COVID-19-positive patients, we included those who had positive polymerase-chain-reaction (PCR), serology, or antigen tests, diagnoses of COVID-19, or diagnosis of post-acute sequelae of SARS-CoV-2 (PASC). For COVID-19-negative patients, we included patients who had no documented SARS-CoV-2 infection and had at least one negative COVID-19 test result within the same study period. A random negative test was chosen as the index date for COVID-19-negative patients. The selection of participants for the COVID-19 positive and negative patient groups in real-world data is summarized in **Figure 1**.

**Figure 1.**
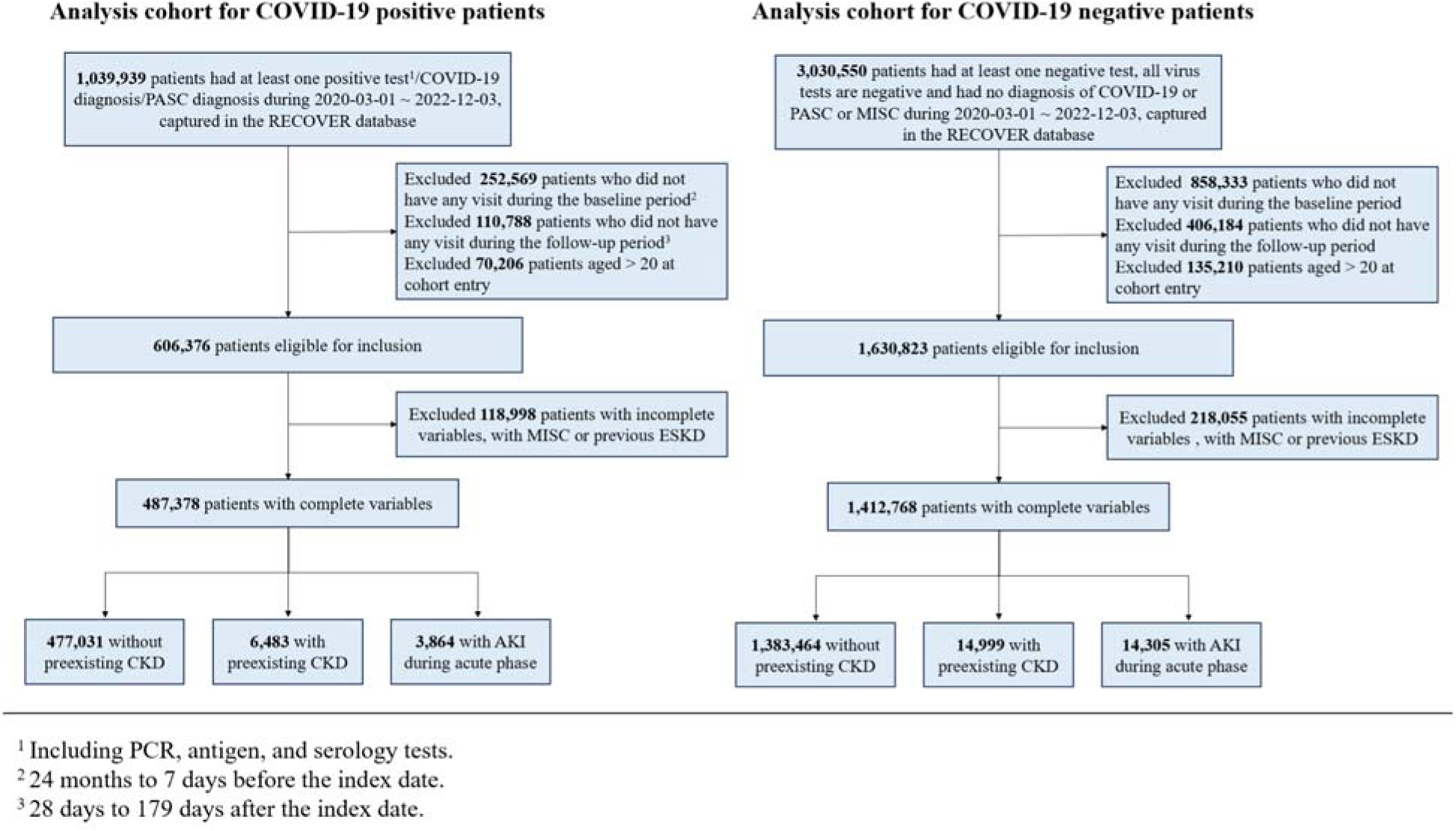
Selection of patients for COVID-19-positive and COVID-19-negative groups.

Patients with either a diagnosis of MIS-C or evidence of end stage kidney disease (ESKD) prior to 28 days after the index date were excluded, where ESKD was defined as having kidney dialysis, kidney transplant, diagnosis of ESKD by ICD-10 codes, or any eGFR ≤ 15 mL/min/1.73 m^2^. This exclusion facilitates a more focused analysis on the direct impact of SARS-CoV-2 on kidney health in the pediatric population^15,16^. We conduct a sensitivity analysis where we do not exclude patients with kidney dialysis during acute phase for the AKI group.

### Kidney Disease Status and Outcome Definitions

Onset date for chronic kidney disease (CKD) stage 2 and higher (2+) was defined as the date of the second eGFR of the earliest pair of eGFR values <90 mL/min/1.73m^2^, separated by >=90 days, without an intervening eGFR >=90. Onset date for CKD stage 3 and higher (3+) was defined as the date of the second eGFR of the earliest pair of eGFR values <60 mL/min/1.73m^2^, separated by >=90 days, without an intervening eGFR >=90. Additionally, we also included two variations to the above definitions by either requiring the eGFR values never return back to above 90 (for CKD stage 2+ and CKD stage 3+) or 60 (for CKD stage 3+), or requiring the majority of eGFR values never return to above 90 or 60. Note that for the definition of CKD outcomes, we require the date of the first eGFR of the pair to be after 28 days of the infection. AKI was defined as having a serum creatinine measurement within 30 days after index date that was 0.3 mg/dl or 50% greater than baseline, where baseline was defined as the closest value within 90 days of index date and if this was not available, established age-/sex-reference values were used^17^. We used the U25 serum creatinine-based equations^18^ to estimate eGFR, due to limited availability of serum cystatin C data. Age was computed at the date of the serum creatinine measurement and the closest height within 180 days of the date of the serum creatinine measurement was used.

For patients without CKD stage 2+ in the 24 months before the SARS-CoV-2 infection, we examined the outcomes of: new onset of CKD stage 2+ or CKD stage 3+ within 28 days to 729 days after SARS-CoV-2 infection. For patients with pre-existing CKD in the 24 months before the SARS-CoV-2 infection, we examined a composite outcome of chronic kidney dialysis, kidney transplant, eGFR decline of ≥50%, ESKD diagnosis, or eGFR ≤15 mL/min/1.73m^2^ within 28 days to 179 days and within 180 days to 729 days after SARS-CoV-2 infection. We also looked at eGFR decline of >=30%, >=40%, and >=50% from within 28 days to 179 days and within 180 days to 729 days after SARS-CoV-2 infection. For patients with AKI during the acute phase (within 28 days) of SARS-CoV-2 infection, we examined the same sets of outcomes for patients in the other two strata and focused on ascertainment of these outcomes within 90 days to 179 days and within 180 days to 729 days.

### Covariates

We examined a comprehensive set of patient characteristics as potential confounders. These included patient age at cohort entry; gender (female, male); race and ethnicity (Asian American/Pacific Islander (AAPI), Hispanic, Non-Hispanic Black (NHB), Non-Hispanic White (NHW), Multiple, Other/Unknown); year-month of cohort entry (from March 2020 to May 2023); site index for cohort entry; obesity defined using age/sex 95th percentile or greater of BMI; a chronic condition indicator as defined by the Pediatric Medical Complexity Algorithm^19^ (PMCA, no chronic condition, non-complex chronic condition, complex chronic condition); a list of pre-existing chronic conditions; the number of inpatient, outpatient, and emergency department (ED) visits; the number of unique medications or prescriptions (0, 1, 2, >=3) at baseline; the number of negative tests 18 months to 7 days before cohort entry date (0, 1, 2, >=3); the number of vaccine doses before cohort entry date (0, 1, >=2); and interval since the last immunization date (no vaccine, <4 months, >=4 months).

### Statistical Analysis

We calculated the incidence of post-acute kidney outcomes in both COVID-19-positive and COVID-19-negative cohorts stratified by pre-existing kidney health status. To quantify the risks of post-acute kidney manifestations of SARS-CoV-2 infection, we used the hazard ratio (HR) as the comparative measure, employing Cox proportional hazards models.

To eliminate the impact of potential measured confounders, we used a propensity score stratification technique with the covariates detailed above. To mitigate the impact of aggregation bias and account for potential clustering arising from site-specific variations, our stratification process required the formation of two comparison groups according to their respective site indices. After performing the stratification, we assessed the standardized mean difference (SMD) between each covariate value for COVID-19 positive and negative patients, with a difference of 0.1 or less indicating an acceptable balance^20,21^. The clinical equipoise before and after matching and SMD of covariates before and after matching can be found in Supplementary Materials Figures S1 and S2.

## Results

### Cohort Identification

A total of 439,182 children and adolescents with COVID-19 and 1,291,075 children and adolescents without COVID-19 in the RECOVER Database were included to evaluate the risks of post-acute kidney manifestations of SARS-CoV-2 infection. Among these patients, 837,558 (50.5%) were male. Baseline comorbidities by COVID-19 status and pre-existing kidney health status are presented in **Table 1**.

**Table 1.** Baseline characteristics of patients, stratified by COVID-19 status and pre-existing kidney health status (acute kidney injury during acute phase of COVID-19 infection (AKI), chronic kidney disease stage 2+ (CKD), and no AKI or CKD).

|  | *COVID-19 Negative* |  |  |  | *COVID-19 Positive* |  |  |  |
| --- | --- | --- | --- | --- | --- | --- | --- | --- |
|  | No AKI or CKD<br>(N=1383464) | CKD<br>(N=14999) | AKI<br>(N=14305) | Overall<br>(N=1412768) | No AKI or CKD<br>(N=477031) | CKD<br>(N=6483) | AKI<br>(N=3864) | Overall<br>(N=487378) |
| **Age at Cohort Entry, year** |  |  |  |  |  |  |  |  |
| **0** | 129940 (9.4%) | 311 (2.1%) | 2714 (19.0%) | 132965 (9.4%) | 50881 (10.7%) | 78 (1.2%) | 394 (10.2%) | 51353 (10.5%) |
| **1** | 121994 (8.8%) | 382 (2.5%) | 975 (6.8%) | 123351 (8.7%) | 36658 (7.7%) | 186 (2.9%) | 263 (6.8%) | 37107 (7.6%) |
| **2** | 96371 (7.0%) | 447 (3.0%) | 789 (5.5%) | 97607 (6.9%) | 27017 (5.7%) | 184 (2.8%) | 200 (5.2%) | 27401 (5.6%) |
| **3** | 88968 (6.4%) | 403 (2.7%) | 626 (4.4%) | 89997 (6.4%) | 23621 (5.0%) | 146 (2.3%) | 174 (4.5%) | 23941 (4.9%) |
| **4** | 84248 (6.1%) | 456 (3.0%) | 465 (3.3%) | 85169 (6.0%) | 22657 (4.7%) | 156 (2.4%) | 126 (3.3%) | 22939 (4.7%) |
| **5** | 81769 (5.9%) | 488 (3.3%) | 480 (3.4%) | 82737 (5.9%) | 20644 (4.3%) | 150 (2.3%) | 120 (3.1%) | 20914 (4.3%) |
| **6** | 72583 (5.2%) | 434 (2.9%) | 472 (3.3%) | 73489 (5.2%) | 19689 (4.1%) | 146 (2.3%) | 119 (3.1%) | 19954 (4.1%) |
| **7** | 64123 (4.6%) | 405 (2.7%) | 393 (2.7%) | 64921 (4.6%) | 19318 (4.0%) | 137 (2.1%) | 97 (2.5%) | 19552 (4.0%) |
| **8** | 58493 (4.2%) | 431 (2.9%) | 356 (2.5%) | 59280 (4.2%) | 19061 (4.0%) | 148 (2.3%) | 102 (2.6%) | 19311 (4.0%) |
| **9** | 54729 (4.0%) | 432 (2.9%) | 366 (2.6%) | 55527 (3.9%) | 19486 (4.1%) | 157 (2.4%) | 102 (2.6%) | 19745 (4.1%) |
| **10** | 51905 (3.8%) | 493 (3.3%) | 392 (2.7%) | 52790 (3.7%) | 20048 (4.2%) | 197 (3.0%) | 106 (2.7%) | 20351 (4.2%) |
| **11** | 51140 (3.7%) | 490 (3.3%) | 360 (2.5%) | 51990 (3.7%) | 21013 (4.4%) | 203 (3.1%) | 76 (2.0%) | 21292 (4.4%) |
| **12** | 50758 (3.7%) | 578 (3.9%) | 444 (3.1%) | 51780 (3.7%) | 19274 (4.0%) | 215 (3.3%) | 108 (2.8%) | 19597 (4.0%) |
| **13** | 51428 (3.7%) | 661 (4.4%) | 577 (4.0%) | 52666 (3.7%) | 20333 (4.3%) | 265 (4.1%) | 130 (3.4%) | 20728 (4.3%) |
| **14** | 52472 (3.8%) | 826 (5.5%) | 635 (4.4%) | 53933 (3.8%) | 21398 (4.5%) | 366 (5.6%) | 180 (4.7%) | 21944 (4.5%) |
| **15** | 53378 (3.9%) | 1099 (7.3%) | 760 (5.3%) | 55237 (3.9%) | 22009 (4.6%) | 499 (7.7%) | 235 (6.1%) | 22743 (4.7%) |
| **16** | 53213 (3.8%) | 1409 (9.4%) | 812 (5.7%) | 55434 (3.9%) | 22286 (4.7%) | 608 (9.4%) | 247 (6.4%) | 23141 (4.7%) |
| **17** | 52051 (3.8%) | 1543 (10.3%) | 919 (6.4%) | 54513 (3.9%) | 21841 (4.6%) | 732 (11.3%) | 300 (7.8%) | 22873 (4.7%) |
| **18** | 42630 (3.1%) | 1335 (8.9%) | 644 (4.5%) | 44609 (3.2%) | 17323 (3.6%) | 641 (9.9%) | 218 (5.6%) | 18182 (3.7%) |
| **19** | 37173 (2.7%) | 1135 (7.6%) | 581 (4.1%) | 38889 (2.8%) | 16653 (3.5%) | 604 (9.3%) | 295 (7.6%) | 17552 (3.6%) |
| **20** | 34098 (2.5%) | 1241 (8.3%) | 545 (3.8%) | 35884 (2.5%) | 15821 (3.3%) | 665 (10.3%) | 272 (7.0%) | 16758 (3.4%) |
| **Sex** |  |  |  |  |  |  |  |  |
| **Female** | 671071 (48.5%) | 8963 (59.8%) | 6477 (45.3%) | 686511 (48.6%) | 237969 (49.9%) | 4052 (62.5%) | 1677 (43.4%) | 243698 (50.0%) |
| **Male** | 712393 (51.5%) | 6036 (40.2%) | 7828 (54.7%) | 726257 (51.4%) | 239062 (50.1%) | 2431 (37.5%) | 2187 (56.6%) | 243680 (50.0%) |
| **Race/ Ethnicity** |  |  |  |  |  |  |  |  |
| **Asian Americans and Pacific Islanders** | 67455 (4.9%) | 554 (3.7%) | 624 (4.4%) | 68633 (4.9%) | 22661 (4.8%) | 193 (3.0%) | 141 (3.6%) | 22995 (4.7%) |
| **Non-Hispanic Black** | 234968 (17.0%) | 3058 (20.4%) | 4045 (28.3%) | 242071 (17.1%) | 83295 (17.5%) | 1478 (22.8%) | 1158 (30.0%) | 85931 (17.6%) |
| **Hispanic** | 285725<br>(20.7%) | 2234<br>(14.9%) | 2673<br>(18.7%) | 290632<br>(20.6%) | 106979<br>(22.4%) | 969<br>(14.9%) | 727<br>(18.8%) | 108675<br>(22.3%) |
| **Multiple** | 33778 (2.4%) | 252 (1.7%) | 164 (1.1%) | 34194 (2.4%) | 9479 (2.0%) | 79 (1.2%) | 38 (1.0%) | 9596 (2.0%) |
| **Other/Unknown** | 135917 (9.8%) | 810 (5.4%) | 1074<br>(7.5%) | 137801 (9.8%) | 46371 (9.7%) | 286<br>(4.4%) | 238<br>(6.2%) | 46895 (9.6%) |
| **Non-Hispanic White** | 625621<br>(45.2%) | 8091<br>(53.9%) | 5725<br>(40.0%) | 639437<br>(45.3%) | 208246<br>(43.7%) | 3478<br>(53.6%) | 1562<br>(40.4%) | 213286<br>(43.8%) |
| **Site** |  |  |  |  |  |  |  |  |
| **A** | 109645 (7.9%) | 633 (4.2%) | 449 (3.1%) | 110727 (7.8%) | 32798 (6.9%) | 272<br>(4.2%) | 140<br>(3.6%) | 33210 (6.8%) |
| **B** | 151321<br>(10.9%) | 1262<br>(8.4%) | 779 (5.4%) | 153362<br>(10.9%) | 55206<br>(11.6%) | 468<br>(7.2%) | 187<br>(4.8%) | 55861<br>(11.5%) |
| **C** | 84313 (6.1%) | 816 (5.4%) | 456 (3.2%) | 85585 (6.1%) | 18541 (3.9%) | 227<br>(3.5%) | 120<br>(3.1%) | 18888 (3.9%) |
| **D** | 44458 (3.2%) | 879 (5.9%) | 467 (3.3%) | 45804 (3.2%) | 16023 (3.4%) | 377<br>(5.8%) | 131<br>(3.4%) | 16531 (3.4%) |
| **E** | 421 (0.0%) | 3 (0.0%) | 5 (0.0%) | 429 (0.0%) | 95 (0.0%) | 0 (0%) | 1 (0.0%) | 96 (0.0%) |
| **F** | 29699 (2.1%) | 273 (1.8%) | 179 (1.3%) | 30151 (2.1%) | 14737 (3.1%) | 140<br>(2.2%) | 68 (1.8%) | 14945 (3.1%) |
| **G** | 530 (0.0%) | 26 (0.2%) | 6 (0.0%) | 562 (0.0%) | 110 (0.0%) | 2 (0.0%) | 1 (0.0%) | 113 (0.0%) |
| **H** | 42704 (3.1%) | 439 (2.9%) | 320 (2.2%) | 43463 (3.1%) | 8618 (1.8%) | 169<br>(2.6%) | 77 (2.0%) | 8864 (1.8%) |
| **I** | 15476 (1.1%) | 141 (0.9%) | 60 (0.4%) | 15677 (1.1%) | 7593 (1.6%) | 76 (1.2%) | 43 (1.1%) | 7712 (1.6%) |
| **J** | 59857 (4.3%) | 1204<br>(8.0%) | 498 (3.5%) | 61559 (4.4%) | 14766 (3.1%) | 381<br>(5.9%) | 101<br>(2.6%) | 15248 (3.1%) |
| **K** | 19641 (1.4%) | 236 (1.6%) | 113 (0.8%) | 19990 (1.4%) | 8605 (1.8%) | 180<br>(2.8%) | 53 (1.4%) | 8838 (1.8%) |
| **L** | 36718 (2.7%) | 271 (1.8%) | 4 (0.0%) | 36993 (2.6%) | 9766 (2.0%) | 61 (0.9%) | 3 (0.1%) | 9830 (2.0%) |
| **M** | 19207 (1.4%) | 809 (5.4%) | 155 (1.1%) | 20171 (1.4%) | 6815 (1.4%) | 260<br>(4.0%) | 50 (1.3%) | 7125 (1.5%) |
| **N** | 98646 (7.1%) | 55 (0.4%) | 5852<br>(40.9%) | 104553 (7.4%) | 41773 (8.8%) | 31 (0.5%) | 1115<br>(28.9%) | 42919 (8.8%) |
| **O** | 122426 (8.8%) | 668 (4.5%) | 487 (3.4%) | 123581 (8.7%) | 27068 (5.7%) | 206<br>(3.2%) | 167<br>(4.3%) | 27441 (5.6%) |
| **P** | 6066 (0.4%) | 133 (0.9%) | 79 (0.6%) | 6278 (0.4%) | 2868 (0.6%) | 68 (1.0%) | 26 (0.7%) | 2962 (0.6%) |
| **Q** | 97556 (7.1%) | 1014<br>(6.8%) | 358 (2.5%) | 98928 (7.0%) | 25771 (5.4%) | 310<br>(4.8%) | 131<br>(3.4%) | 26212 (5.4%) |
| **R** | 51734 (3.7%) | 684 (4.6%) | 253 (1.8%) | 52671 (3.7%) | 20695 (4.3%) | 385<br>(5.9%) | 80 (2.1%) | 21160 (4.3%) |
| **S** | 35663 (2.6%) | 1165<br>(7.8%) | 442 (3.1%) | 37270 (2.6%) | 10139 (2.1%) | 435<br>(6.7%) | 132<br>(3.4%) | 10706 (2.2%) |
| **T** | 135538 (9.8%) | 179 (1.2%) | 68 (0.5%) | 135785 (9.6%) | 63376<br>(13.3%) | 127<br>(2.0%) | 31 (0.8%) | 63534<br>(13.0%) |
| **U** | 2318 (0.2%) | 1 (0.0%) | 0 (0%) | 2319 (0.2%) | 857 (0.2%) | 1 (0.0%) | 0 (0%) | 858 (0.2%) |
| **V** | 15410 (1.1%) | 164 (1.1%) | 120 (0.8%) | 15694 (1.1%) | 27489 (5.8%) | 481<br>(7.4%) | 194<br>(5.0%) | 28164 (5.8%) |
| **W** | 30144 (2.2%) | 211 (1.4%) | 144 (1.0%) | 30499 (2.2%) | 4963 (1.0%) | 68 (1.0%) | 52 (1.3%) | 5083 (1.0%) |
| **X** | 43830 (3.2%) | 206 (1.4%) | 349 (2.4%) | 44385 (3.1%) | 13302 (2.8%) | 93 (1.4%) | 74 (1.9%) | 13469 (2.8%) |
| **Y** | 45210 (3.3%) | 453 (3.0%) | 338 (2.4%) | 46001 (3.3%) | 12945 (2.7%) | 161<br>(2.5%) | 101<br>(2.6%) | 13207 (2.7%) |
| **Z** | 35747 (2.6%) | 2243<br>(15.0%) | 1860<br>(13.0%) | 39850 (2.8%) | 13197 (2.8%) | 1106<br>(17.1%) | 629<br>(16.3%) | 14932 (3.1%) |
| **AA** | 32692 (2.4%) | 600 (4.0%) | 166 (1.2%) | 33458 (2.4%) | 12870 (2.7%) | 330<br>(5.1%) | 67 (1.7%) | 13267 (2.7%) |
| **BB** | 16494 (1.2%) | 231 (1.5%) | 298 (2.1%) | 17023 (1.2%) | 6045 (1.3%) | 68 (1.0%) | 90 (2.3%) | 6203 (1.3%) |
| **Cohort Entry Period** |  |  |  |  |  |  |  |  |
| **03/2020 - 05/2020** | 25471 (1.8%) | 594 (4.0%) | 436 (3.0%) | 26501 (1.9%) | 4778 (1.0%) | 96 (1.5%) | 114<br>(3.0%) | 4988 (1.0%) |
| **03/2021 - 05/2021** | 137964<br>(10.0%) | 1497<br>(10.0%) | 1534<br>(10.7%) | 140995<br>(10.0%) | 28730 (6.0%) | 448<br>(6.9%) | 280<br>(7.2%) | 29458 (6.0%) |
| **03/2022 - 05/2022** | 114323 (8.3%) | 1110<br>(7.4%) | 1401<br>(9.8%) | 116834 (8.3%) | 36985 (7.8%) | 509<br>(7.9%) | 337<br>(8.7%) | 37831 (7.8%) |
| **06/2020 - 08/2020** | 96457 (7.0%) | 1731<br>(11.5%) | 938 (6.6%) | 99126 (7.0%) | 17368 (3.6%) | 248<br>(3.8%) | 169<br>(4.4%) | 17785 (3.6%) |
| **06/2021 - 08/2021** | 135823 (9.8%) | 1431<br>(9.5%) | 1477<br>(10.3%) | 138731 (9.8%) | 26162 (5.5%) | 409<br>(6.3%) | 268<br>(6.9%) | 26839 (5.5%) |
| **06/2022 - 08/2022** | 83688 (6.0%) | 958 (6.4%) | 1336 (9.3%) | 85982 (6.1%) | 51659 (10.8%) | 757 (11.7%) | 456 (11.8%) | 52872 (10.8%) |
| **09/2020 - 11/2020** | 140935 (10.2%) | 1798 (12.0%) | 1506 (10.5%) | 144239 (10.2%) | 32030 (6.7%) | 408 (6.3%) | 224 (5.8%) | 32662 (6.7%) |
| **09/2021 - 11/2021** | 217656 (15.7%) | 1645 (11.0%) | 1556 (10.9%) | 220857 (15.6%) | 49458 (10.4%) | 630 (9.7%) | 409 (10.6%) | 50497 (10.4%) |
| **09/2022 - 12/2022** | 132831 (9.6%) | 1216 (8.1%) | 1393 (9.7%) | 135440 (9.6%) | 26934 (5.6%) | 422 (6.5%) | 272 (7.0%) | 27628 (5.7%) |
| **12/2020 - 02/2021** | 135978 (9.8%) | 1673 (11.2%) | 1341 (9.4%) | 138992 (9.8%) | 51601 (10.8%) | 665 (10.3%) | 356 (9.2%) | 52622 (10.8%) |
| **12/2021 - 02/2022** | 162338 (11.7%) | 1346 (9.0%) | 1387 (9.7%) | 165071 (11.7%) | 151326 (31.7%) | 1891 (29.2%) | 979 (25.3%) | 154196 (31.6%) |
| **Obesity** |  |  |  |  |  |  |  |  |
| **Non-Obese** | 722415 (52.2%) | 7439 (49.6%) | 7852 (54.9%) | 737706 (52.2%) | 214869 (45.0%) | 2723 (42.0%) | 1910 (49.4%) | 219502 (45.0%) |
| **Obese** | 497849 (36.0%) | 7541 (50.3%) | 4836 (33.8%) | 510226 (36.1%) | 206568 (43.3%) | 3759 (58.0%) | 1567 (40.6%) | 211894 (43.5%) |
| **Unknown** | 163200 (11.8%) | 19 (0.1%) | 1617 (11.3%) | 164836 (11.7%) | 55594 (11.7%) | 1 (0.0%) | 387 (10.0%) | 55982 (11.5%) |
| **Chronic Disease Status** |  |  |  |  |  |  |  |  |
| **0** | 1006146 (72.7%) | 2840 (18.9%) | 6272 (43.8%) | 1015258 (71.9%) | 351288 (73.6%) | 1283 (19.8%) | 1641 (42.5%) | 354212 (72.7%) |
| **1** | 220119 (15.9%) | 3280 (21.9%) | 2400 (16.8%) | 225799 (16.0%) | 75835 (15.9%) | 1403 (21.6%) | 585 (15.1%) | 77823 (16.0%) |
| **2** | 157199 (11.4%) | 8879 (59.2%) | 5633 (39.4%) | 171711 (12.2%) | 49908 (10.5%) | 3797 (58.6%) | 1638 (42.4%) | 55343 (11.4%) |
| **Number of Tests** |  |  |  |  |  |  |  |  |
| **0** | 1049258 (75.8%) | 9026 (60.2%) | 9148 (63.9%) | 1067432 (75.6%) | 291958 (61.2%) | 2415 (37.3%) | 1916 (49.6%) | 296289 (60.8%) |
| **1** | 218747 (15.8%) | 2818 (18.8%) | 2534 (17.7%) | 224099 (15.9%) | 101336 (21.2%) | 1459 (22.5%) | 709 (18.3%) | 103504 (21.2%) |
| **2** | 66084 (4.8%) | 1316 (8.8%) | 1059 (7.4%) | 68459 (4.8%) | 41027 (8.6%) | 844 (13.0%) | 360 (9.3%) | 42231 (8.7%) |
| **>2** | 49375 (3.6%) | 1839 (12.3%) | 1564 (10.9%) | 52778 (3.7%) | 42710 (9.0%) | 1765 (27.2%) | 879 (22.7%) | 45354 (9.3%) |
| **Vaccine dosage** |  |  |  |  |  |  |  |  |
| **0** | 1248073 (90.2%) | 12690 (84.6%) | 13165 (92.0%) | 1273928 (90.2%) | 416466 (87.3%) | 5189 (80.0%) | 3399 (88.0%) | 425054 (87.2%) |
| **1** | 25661 (1.9%) | 417 (2.8%) | 260 (1.8%) | 26338 (1.9%) | 12185 (2.6%) | 288 (4.4%) | 135 (3.5%) | 12608 (2.6%) |
| **>=2** | 109730 (7.9%) | 1892 (12.6%) | 880 (6.2%) | 112502 (8.0%) | 48380 (10.1%) | 1006 (15.5%) | 330 (8.5%) | 49716 (10.2%) |
| **Number of Drugs** |  |  |  |  |  |  |  |  |
| **0** | 379837 (27.5%) | 355 (2.4%) | 2029 (14.2%) | 382221 (27.1%) | 117622 (24.7%) | 124 (1.9%) | 529 (13.7%) | 118275 (24.3%) |
| **1** | 173275 (12.5%) | 393 (2.6%) | 1040 (7.3%) | 174708 (12.4%) | 59593 (12.5%) | 182 (2.8%) | 258 (6.7%) | 60033 (12.3%) |
| **2** | 136222 (9.8%) | 490 (3.3%) | 898 (6.3%) | 137610 (9.7%) | 49144 (10.3%) | 181 (2.8%) | 228 (5.9%) | 49553 (10.2%) |
| **>2** | 694130 (50.2%) | 13761 (91.7%) | 10338 (72.3%) | 718229 (50.8%) | 250672 (52.5%) | 5996 (92.5%) | 2849 (73.7%) | 259517 (53.2%) |

### Incidence and risk of post-acute kidney outcomes for COVID-19-positive and COVID-19-negative patients

**Table 2** presents the incidence of individual and composite post-acute kidney outcomes in the COVID-19-positive cohort compared with the COVID-19-negative cohort, stratified by pre-existing kidney health status. Notably, for kidney outcomes within the post-acute phase, the incidence in COVID-19-positive patients was higher than that in COVID-19-negative patients for all outcomes across all three strata.

**Table 2.** Raw incidence (in %) of individual and composite kidney outcomes, stratified by COVID-19 status and pre-existing kidney health status (acute kidney injury during acute phase of COVID-19 infection (AKI), chronic kidney disease stage 2+ (CKD), and no AKI or CKD).

|  |  | Post-acute phase (%) (days 90-179) |  | Chronic phase (%) (days 180-729) |  |
| --- | --- | --- | --- | --- | --- |
| AKI |  | COVID-19 (n=3864) | Control (n=14305) | COVID-19 (n=3864) | Control (n=14305) |
|  | Composite outcome | 5.33% | 3.33% | 9.58% | 5.56% |
|  | eGFR decline of 50% or more | 2.28% | 1.34% | 4.19% | 2.15% |
|  | eGFR decline of 40% or more | 3.34% | 2.18% | 6.63% | 3.61% |
|  | eGFR decline of 30% or more | 5.25% | 3.33% | 9.45% | 5.56% |
|  |  | Post-acute phase (%) (days 28-729) |  |  |  |
|  | CKD 2+ | 5.02% | 3.53% |  |  |
|  | CKD 2+ (majority not returned to 90) | 4.52% | 3.09% |  |  |
|  | CKD 2+ (not returned to 90) | 4.12% | 2.79% |  |  |
|  | CKD 3+ | 1.61% | 0.82% |  |  |
|  | CKD 3+ (majority not returned to 90) | 1.37% | 0.65% |  |  |
|  | CKD 3+ (not returned to 90) | 1.53% | 0.73% |  |  |
|  | CKD 3+ (not returned to 60) | 1.29% | 0.61% |  |  |
|  |  | Post-acute phase (%) (days 28-179) |  | Chronic phase (%) (days 180-729) |  |
| CKD |  | COVID-19 (n=6483) | Control (n=14999) | COVID-19 (n=6483) | Control (n=14999) |
|  | Composite outcome | 13.388% | 10.932% | 15.876% | 15.653% |
|  | eGFR decline of 50% or more | 3.283% | 3.023% | 4.227% | 4.385% |
|  | eGFR decline of 40% or more | 7.552% | 6.301% | 8.796% | 8.998% |
|  | eGFR decline of 30% or more | 13.152% | 10.932% | 15.619% | 15.653% |
|  |  | Post-acute phase (%) (days 28-729) |  |  |  |
| No AKI or CKD |  | COVID-19 (n=477031) | Control (n=1383464) |  |  |
|  | CKD 2+ | 0.329% | 0.282% |  |  |
|  | CKD 2+ (majority not returned to 90 and above) | 0.284% | 0.236% |  |  |
|  | CKD 2+ (not returned to 90 and above) | 0.262% | 0.210% |  |  |
|  | CKD 3+ | 0.028% | 0.022% |  |  |
|  | CKD 3+ (majority not returned to 90 and above) | 0.021% | 0.016% |  |  |
|  | CKD 3+ (not returned to 90 and above) | 0.023% | 0.020% |  |  |
|  | CKD level 3 & above (not returned to 60 and above) | 0.018% | 0.014% |  |  |

**Figure 2** shows the risks of post-acute kidney manifestations of SARS-CoV-2 infection, stratified by pre-existing kidney health status. For patients without pre-existing CKD, our findings indicate statistically significant increased risks in several post-acute kidney outcomes following SARS-CoV-2 infection. The risk of new onset of CKD stage 2+ was significantly increased during the post-acute phase (days 28-729) with a HR of 1.17 (95% CI: 1.12-1.22), and the HR for CKD stage 2+ with eGFR never returning to 90 mL/min/1.73m^2^ and above was 1.20 (95% CI: 1.13-1.28). For the outcome of CKD stage 3+ during days 28-729, the HR was 1.35 (95% CI: 1.13-1.62), and for CKD 3+ without return of eGFR to 60 mL/min/1.73m^2^ and above, the HR was 1.35 (95% CI: 1.15-1.59). These significant findings underscore the impact of SARS-CoV-2 on long-term kidney health.

**Figure 2.**
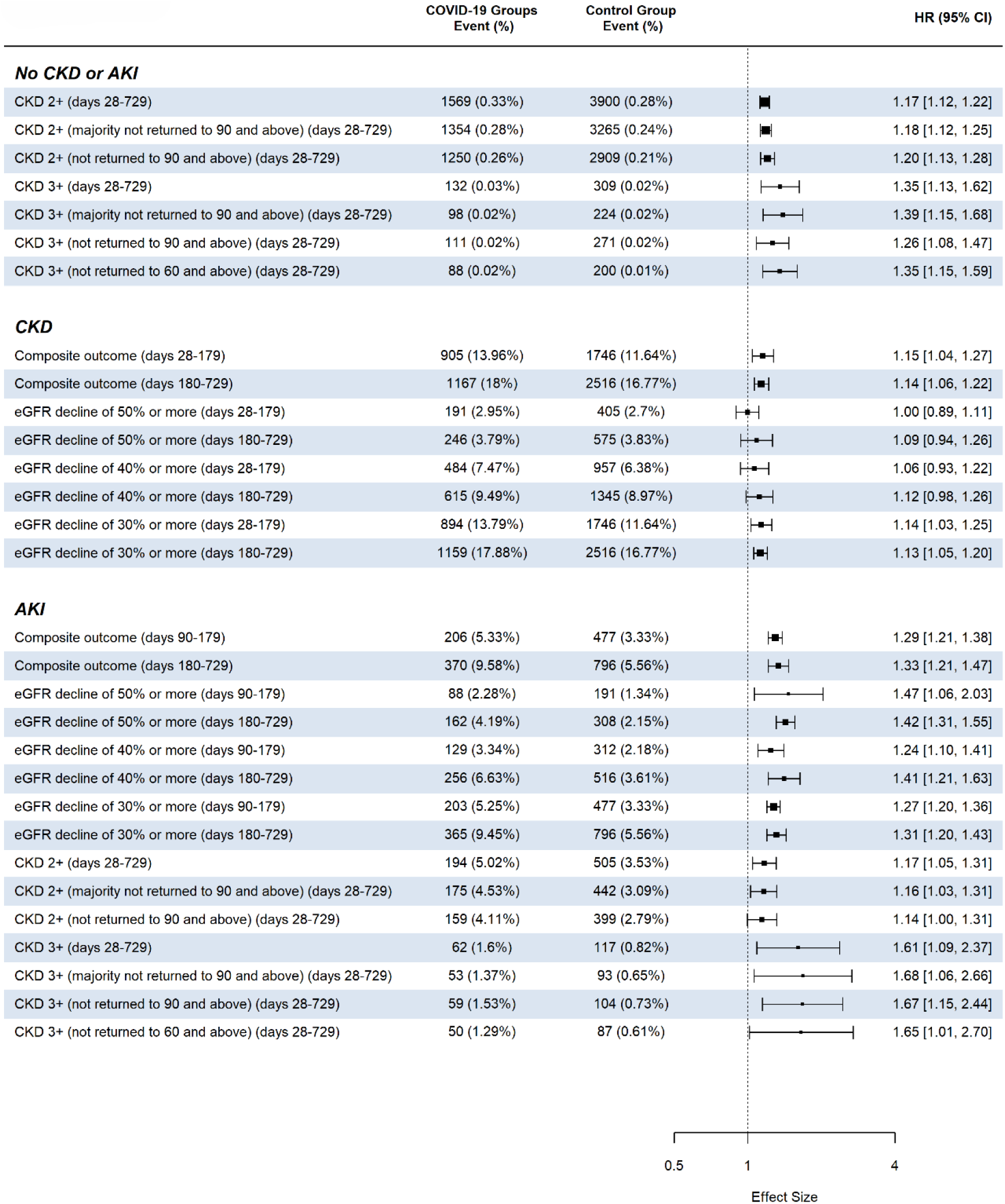
Adjusted hazard ratios for the post-acute phase and the chronic phase comparing COVID-19 positive patients to COVID-19 negative patients based on pre-existing kidney function status (acute kidney injury during acute phase of COVID-19 infection (AKI), chronic kidney disease stage 2+ (CKD), and no AKI or CKD).

For patients with pre-existing CKD, we observed statistically significant increased risks post SARS-CoV-2 infection in several outcomes. Specifically, there was an increased risk of the composite kidney outcome between days 28-179, with an HR of 1.15 (95% CI: 1.04-1.27), as well as an HR of 1.14 (95% CI: 1.06-1.22) for days 180-729. Additionally, an increase in risk was found for an eGFR decline of >=30% between days 28-179, with a HR of 1.14 (95% CI: 1.03-1.25), and an eGFR decline of >=30% between days 180-729, with a HR of 1.13 (95% CI: 1.05-1.20). These findings indicate a heightened risk for kidney function decline in individuals with pre-existing CKD following COVID-19 infection.

For patients who experienced AKI during the acute phase of SARS-CoV-2 infection, the study results indicate statistically significant increased risks during the post-acute phases. The HR for the composite kidney outcome was 1.29 (95% CI: 1.21-1.38) for days 90-179 and 1.33 (95% CI: 1.21-1.47) for days 180-729. There was a significant risk for an eGFR decline of >=50% in both the earlier post-acute phase (HR 1.47; 95% CI: 1.06-2.03) and the later post-acute phase (HR 1.42; 95% CI: 1.31-1.55). There was a significant risk for an eGFR decline of >=40% in both the earlier post-acute phase (HR 1.24; 95% CI: 1.10-1.41) and the later post-acute phase (HR 1.41; 95% CI: 1.21-1.63). Additionally, a significant risk was observed for an eGFR decline of >=30% at 1.27 (95% CI: 1.20-1.36) for days 28-179 and remained high at 1.31 (95% CI: 1.20-1.43) for days 180-729. These findings highlight a sustained increased risk for adverse kidney outcomes in patients with AKI post-COVID-19.

## Discussion

### Principal Findings

In this population-based study, using a cohort of 1,864,637 patients under the age of 21 in the United States, our results reveal a significant increase in the risk of various kidney outcomes associated with COVID-19 infection. This heightened risk includes a new onset of mild-moderate CKD during the post-acute phase of the infection. For patients with pre-existing CKD and patients who experienced AKI during the acute phase, we observed an increased risk of a composite outcome of >=50% eGFR decline, eGFR ≤15 mL/min/1.73m^2^, dialysis, kidney transplant, or ESKD diagnosis.

### Comparison with Other Studies

Our findings align with prior research indicating that COVID-19 infection elevates the risk of kidney outcomes. Many of these earlier studies were limited by insufficient sample sizes for precise estimations and restricted their risk assessments to specific outcomes, such as AKI, and time frames^7,11–14^. Our results are also consistent with a study that utilized the US Department of Veterans Affairs national health care databases (n=1,726,683) to examine risk of adverse kidney outcomes in the post-acute phase following infection^10^. Bowe et al.^10^ observed a significantly higher risk of adverse kidney outcomes among COVID-19-infected individuals, including eGFR decline ≥30% (aHR 1.25; 95% CI: 1.14 to 1.37), eGFR decline ≥40% (aHR 1.44; 95% CI: 1.37 to 1.51), eGFR decline ≥ 50% (aHR 1.62; 95% CI: 1.51 to 1.74), and ESKD (aHR, 2.96; 95% CI, 2.49 to 3.51). Our study uniquely focused on children and adolescents, extending the evaluation of risk into the chronic phase beyond the post-acute period. We also included additional outcomes of new onset CKD. In addition, we stratified the patients based on their pre-existing kidney function status to examine the impact of PASC on adverse kidney outcomes.

### Interpretation

The precise mechanisms underlying the observed associations in our study remain unclear. One plausible explanation could be attributed to the direct impact of COVID-19 on the kidneys, evidenced by the persistence of SARS-CoV-2 in tissues and prolonged virus shedding^22,23^.

Concurrently, the chronic inflammation induced by COVID-19 infection might adversely affect hemodynamic stability, potentially leading to kidney injury^24^. An alternative explanation may lie in the therapeutic interventions employed to manage severe COVID-19 cases^24^, as well as the broader economic and social conditions resulting from the pandemic^25,26^. Further investigations are warranted to elucidate the intricate pathways through which these factors interplay, contributing to the observed associations in our study.

### Limitations and Strengths

Our study has certain limitations. First, while we successfully achieved a balanced distribution of baseline covariates through matching, the propensity scores were constructed using only the variables accessible within our study databases. Nevertheless, it is important to highlight that our findings remained consistent and robust through sensitivity analyses aimed at addressing residual confounding. We conducted sensitivity analyses to stratify patients by age, gender, race, hospitalization status, obesity defined using age/sex 95th percentile or greater, dominant variant periods, and COVID infection severity levels and the results can be found in the supplementary materials.

Second, misclassification might exist for both of AKI and CKD outcomes. For AKI, if there was no baseline info available, we could potentially be classifying as AKI someone with underlying CKD. For long term outcomes, we could potentially capture multiple episodes of AKI as CKD. However, our various definitions of CKD try to mitigate this possibility.

Another potential limitation is that it is difficult to ascertain from this study on whether the increased risk associated with COVID-19 infection in the pre-existing CKD group are inherently due to the COVID-19 infection itself or whether the increased risk reflects a more general risk related to patients with pre-existing CKD who get sick with an illness may see negative impact on eGFR. However, the findings in the AKI group and the No AKI or CKD groups do raise concern that there is an inherent risk to COVID-19 infection itself.

In addition, patients with positive home testing (i.e. with antigen testing available in 2021 and 2022) that would not be accessible/visible in the EHR, could inadvertently be incorporated into the COVID-negative control group. However, this would also serve to bias towards the null hypothesis, and would only strength the conclusion of the increased risk of CKD from post-acute sequelae of COVID. However, patients with hospital-based testing may have been more ill from COVID than those patients testing positive at home, and this could also be a potential bias/limitation of this data.

A strength of this study is its utilization of large, population-based databases within RECOVER, including nineteen children’s hospitals and health institutions in the US. This large program facilitated the precise identification of comprehensive COVID-19 infection records and visits throughout the entire study period in the United States, thereby mitigating the potential for selection bias. Furthermore, using a decentralized COVID-19 infection registry provided detailed and accurate information on infection status, reducing the likelihood of exposure misclassification bias. Notably, our investigation extended into the chronic phase, with a follow-up period of up to two years, representing one of the longest follow-up durations reported for children and adolescents exposed to COVID-19 infection in the United States.

This ongoing research is pivotal not only for improving the clinical management of individuals with persistent kidney outcomes and diseases but also for informing public health strategies aimed at preventing long-term consequences of SARS-CoV-2 infection.

## Data Availability

The data is available through applying through the RECOVER program.

## Disclosure Statement

### Disclaimer

This content is solely the responsibility of the authors and does not necessarily represent the official views of the RECOVER Initiative, the NIH, or other funders.

### Potential Conflicts of Interest

Dr. Jhaveri is a consultant for AstraZeneca, Seqirus, Dynavax, receives an editorial stipend from Elsevier and Pediatric Infectious Diseases Society and royalties from Up To Date/Wolters Kluwer.

Dr. Ruby Patel is the Primary Investigator for FIONA study, no stipend or compensation being given.

Dr. Modi reports research funding outside of this work from the National Institutes of Health, Centers for Disease Control, the Patient-Centered Outcomes Research Institute, Travere Therapeutics, and Boehringer Ingelheim. He is also the current director of the Kidney Research Network Data Coordinating Center.

Dr. Harshman reports research funding outside of this work from the National Institutes of Health.

Dr. Harshman reports research funding outside of this work from Bayer Pharmaceuticals.

Dr. Verghese reports research funding outside of this work from the Department of Defense and Viracor Pharmaceuticals.

Dr. Dixon reports consultancies with Novartis Pharmaceuticals, Alexion Astra Zeneca Rare Disease, Apellis Pharmaceuticals, and Arrowhead Pharmaceuticals.

## Acknowledgements

This study is part of the NIH Researching COVID to Enhance Recovery (RECOVER) Initiative, which seeks to understand, treat, and prevent the post-acute sequelae of SARS-CoV-2 infection (PASC). For more information on RECOVER, visit https://recovercovid.org/

We would like to thank the National Community Engagement Group (NCEG), all patient, caregiver and community Representatives, and all the participants enrolled in the RECOVER Initiative. We would like to thank the patient representatives Maxwell Hornig and Leyna Aragon for their helpful suggestions and comments.

## Data sharing statement

The research data is confidential.

## References

1. Proal AD, VanElzakker MB. Long COVID or Post-acute Sequelae of COVID-19 (PASC): An Overview of Biological Factors That May Contribute to Persistent Symptoms. Front Microbiol. 2021;12:698169. doi:10.3389/fmicb.2021.698169

2. Davis HE, McCorkell L, Vogel JM, Topol EJ. Long COVID: major findings, mechanisms and recommendations. Nat Rev Microbiol. 2023;21(3):133–146. doi:10.1038/s41579-022-00846-2

3. Vahratian A, Adjaye-Gbewonyo D, Lin JMS, Saydah S. Long COVID Among Children: United States, 2022. National Center for Health Statistics (U.S.); 2023. doi:10.15620/cdc:132416

4. Soriano JB, Murthy S, Marshall JC, Relan P, Diaz JV. A clinical case definition of post-COVID-19 condition by a Delphi consensus. The Lancet Infectious Diseases. 2022;22(4):e102–e107. doi:10.1016/S1473-3099(21)00703-9

5. Fang Z, Ahrnsbrak R, Rekito A. Evidence Mounts That About 7% of US Adults Have Had Long COVID. JAMA. Published online June 7, 2024. doi:10.1001/jama.2024.11370

6. Rao S, Gross RS, Mohandas S, et al. Postacute Sequelae of SARS-CoV-2 in Children. Pediatrics. 2024;153(3):e2023062570. doi:10.1542/peds.2023-062570

7. Ludvigsson JF. Systematic review of COVID-19 in children shows milder cases and a better prognosis than adults. Acta Paediatr. 2020;109(6):1088–1095. doi:10.1111/apa.15270

8. Pezzullo AM, Axfors C, Contopoulos-Ioannidis DG, Apostolatos A, Ioannidis JPA. Age-stratified infection fatality rate of COVID-19 in the non-elderly population. Environ Res. 2023;216(Pt 3):114655. doi:10.1016/j.envres.2022.114655

9. Hasnat F, Noman F, Moben AL, Sarker AS, Morshed J, Mutanabbi M. Difference in Clinical Patterns between COVID-19 Affected Children and Adults. Mymensingh Med J. 2021;30(4):1093–1099.

10. Bowe B, Xie Y, Xu E, Al-Aly Z. Kidney Outcomes in Long COVID. Journal of the American Society of Nephrology. 2021;32(11):2851. doi:10.1681/ASN.2021060734

11. Basalely A, Gurusinghe S, Schneider J, et al. Acute kidney injury in pediatric patients hospitalized with acute COVID-19 and multisystem inflammatory syndrome in children associated with COVID-19. Kidney International. 2021;100(1):138–145. doi:10.1016/j.kint.2021.02.026

12. Nomura E, Finn LS, Bauer A, et al. Pathology findings in pediatric patients with COVID-19 and kidney dysfunction. Pediatr Nephrol. 2022;37(10):2375–2381. doi:10.1007/s00467-022-05457-w

13. Raina R, Chakraborty R, Mawby I, Agarwal N, Sethi S, Forbes M. Critical analysis of acute kidney injury in pediatric COVID-19 patients in the intensive care unit. Pediatr Nephrol. 2021;36(9):2627–2638. doi:10.1007/s00467-021-05084-x

14. Serafinelli J, Mastrangelo A, Morello W, et al. Kidney involvement and histological findings in two pediatric COVID-19 patients. Pediatr Nephrol. 2021;36(11):3789–3793. doi:10.1007/s00467-021-05212-7

15. Tripathi AK, Pilania RK, Bhatt GC, Atlani M, Kumar A, Malik S. Acute kidney injury following multisystem inflammatory syndrome associated with SARS-CoV-2 infection in children: a systematic review and meta-analysis. Pediatr Nephrol. 2023;38(2):357–370. doi:10.1007/s00467-022-05701-3

16. Guimarães D, Pissarra R, Reis-Melo A, Guimarães H. Multisystem inflammatory syndrome in children (MISC): A systematic review. International Journal of Clinical Practice. 2021;75(11):e14450. doi:10.1111/ijcp.14450

17. Fitzgerald JC, Basu RK, Fuhrman DY, et al. Renal Dysfunction Criteria in Critically Ill Children: The PODIUM Consensus Conference. Pediatrics. 2022;149(1 Suppl 1):S66–S73. doi:10.1542/peds.2021-052888J

18. Pierce CB, Muñoz A, Ng DK, Warady BA, Furth SL, Schwartz GJ. Age- and sex-dependent clinical equations to estimate glomerular filtration rates in children and young adults with chronic kidney disease. Kidney International. 2021;99(4):948–956. doi:10.1016/j.kint.2020.10.047

19. Simon TD, Haaland W, Hawley K, Lambka K, Mangione-Smith R. Development and Validation of the Pediatric Medical Complexity Algorithm (PMCA) Version 3.0. Acad Pediatr. 2018;18(5):577–580. doi:10.1016/j.acap.2018.02.010

20. Austin PC. Balance diagnostics for comparing the distribution of baseline covariates between treatment groups in propensity-score matched samples. Stat Med. 2009;28(25):3083–3107. doi:10.1002/sim.3697

21. Wu Q, Tong J, Zhang B, et al. Real-World Effectiveness of BNT162b2 Against Infection and Severe Diseases in Children and Adolescents. Ann Intern Med. 2024;177(2):165–176. doi:10.7326/M23-1754

22. Swank Z, Senussi Y, Manickas-Hill Z, et al. Persistent Circulating Severe Acute Respiratory Syndrome Coronavirus 2 Spike Is Associated With Post-acute Coronavirus Disease 2019 Sequelae. Clinical Infectious Diseases. 2023;76(3):e487–e490. doi:10.1093/cid/ciac722

23. Li X, Tam AR, Chu WM, et al. Risk Factors for Slow Viral Decline in COVID-19 Patients during the 2022 Omicron Wave. Viruses. 2022;14(8):1714. doi:10.3390/v14081714

24. Schiffl H, Lang SM. Long-term interplay between COVID-19 and chronic kidney disease. International Urology and Nephrology. 2023;55(8):1977–1984. doi:10.1007/s11255-023-03528-x

25. Townsend E. COVID-19 policies in the UK and consequences for mental health. The Lancet Psychiatry. 2020;7(12):1014–1015. doi:10.1016/S2215-0366(20)30457-0

26. Huybrechts KF, Bateman BT, Pawar A, et al. Maternal and fetal outcomes following exposure to duloxetine in pregnancy: cohort study. BMJ. 2020;368:m237. doi:10.1136/bmj.m237

